# VaccineScout: A Computational Framework for Guiding the Composition and Valency of SARS-CoV-2 Vaccine Updates

**DOI:** 10.64898/2026.11.03.26367217

**Authors:** Pranav Gangwar, Sunreet Khanna, Joel O. Wertheim, Bill Lin, Yatish Turakhia

## Abstract

The continual evolution of SARS-CoV-2 necessitates periodic vaccine updates to maintain immunity against newly emerging variants. However, vaccine design remains largely guided by experimental serological studies, and there is no publicly available computational framework to help systematically prioritize vaccine variants and guide valency selection. Here, we analyze over 14 million SARS-CoV-2 genomes collected globally between 2021 and 2025 and find that over half of spike amino acid sequences (primary vaccine target) observed during the first six months after vaccine selection were already circulating at the time of selection, compared with only 5-10% of sequences during the subsequent six months. These findings provide quantitative support for annual COVID-19 vaccine updates. We further show that circulating variant prevalence and open-source variant fitness predictors can both effectively rank circulating variants by their future prevalence.

Motivated by these findings, we developed VaccineScout, a computational framework that recommends vaccine candidates by minimizing the average spike amino acid substitution distances to the forecasted prevalence of circulating variants. VaccineScout also helps determine vaccine valency by quantifying diminishing returns as additional variants are introduced. Retrospective analyses showed that VaccineScout-recommended candidates either matched U.S. Food and Drug Administration recommendations or had lower average spike sequence distances to future variants and higher fitness. In addition, their spike distances to future variants were comparable to an oracle selection informed by future prevalence, indicating near-optimal performance by this metric. Together, VaccineScout introduces an *in silico* framework for systematically prioritizing SARS-CoV-2 vaccine candidates for costly downstream experimental validation.

## Introduction

SARS-CoV-2 continues to evolve, acquiring mutations that enhance its ability to evade antibodies generated by prior infection and vaccination^1,2^. This ongoing evolution contributes to a substantial disease burden, estimated at tens of billions of dollars annually in the United States alone^3^. To reduce this burden, public health authorities, such as the U.S. Food and Drug Administration (FDA) and WHO’s Technical Advisory Group on COVID-19 Vaccine Composition (TAG-CO-VAC), recommend annual COVID-19 vaccine updates incorporating newly emerging variants^4,5^. These vaccines primarily target the SARS-CoV-2 spike protein, which mediates viral entry into host cells^6,7^, and can be multivalent^8^, i.e., include multiple variants. Updating a vaccine therefore requires two key decisions: which variants to include and how many. The first requires assessing whether existing vaccines remain effective against emerging variants and identifying variants for reformulation when protection wanes. This process relies largely on time-consuming experiments using animal and human sera^9,10^, which cannot be afforded for all possible combinations of circulating variants across different valencies. In addition to experimental evidence, the FDA considers recent genomic surveillance data when recommending vaccine variants, as experiments often lag behind evolving epidemiological trends. This is illustrated by the 2024-2025 season, when rapidly changing epidemiological trends prompted the FDA to revise its recommendation from the JN.1 to the KP.2 variant within days^11,12^. Vaccine valency similarly lacks a systematic framework, with the FDA having recommended both monovalent and bivalent formulations^8,11,13,14^, even as higher-valency vaccines remain feasible to manufacture^15,16^. While some closed-source models^17^ have been developed to predict immune escape from proprietary serum neutralization measurements and Deep Mutational Scanning (DMS) data^18–22^, a systematic *in silico* approach that quickly adapts to genomic surveillance trends and jointly recommends vaccine composition and valency remains lacking.

Several computational approaches have been explored for SARS-CoV-2 vaccine variant selection and design. A closed-source model^17^ predicts immune escape from proprietary serum neutralization measurements and Deep Mutational Scanning (DMS) data^18–22^, but its dependence on these proprietary inputs and its inability to recommend valency limit both accessibility and applicability. Other approaches^23,24^ have attempted to design broadly protective SARS-CoV-2 spike sequences *de novo*, combining known antibody-escape mutations identified through DMS and ACE2 binding data^25–27^. However, these generate artificial spike sequences whose effectiveness depends on whether their constituent mutations arise in future variants. Because these vaccine sequences can be antigenically different from both circulating and future variants, they may elicit weaker immune responses against those variants^24,28^. In comparison, vaccines containing currently circulating variants, including those used by the FDA, may lose effectiveness when a genetically distinct variant becomes dominant^29,30^, but provide stronger protection while the selected circulating variant remains dominant. Consequently, vaccines based on circulating variants remain predominant in practice. A recent influenza (H3N2 and H1N1) vaccine design framework integrates genomic surveillance data with extensive human and animal antisera data to select circulating variants for annual updates^31^. However, applying this approach to SARS-CoV-2 is challenging because antisera data spanning different variants are not publicly available. Several open-source models can accurately predict SARS-CoV-2 variant fitness or evolutionary trajectories from genome sequencing data alone^32–35^, which could potentially substitute for antisera data, but their utility for vaccine design remains unexplored. Consequently, a systematic, publicly available *in silico* method that adapts quickly to genomic surveillance trends and jointly recommends vaccine composition and valency remains lacking.

Here, we present a systematic, large-scale study of the SARS-CoV-2 vaccine design problem. We first estimate how long a vaccine designed with circulating variants remains effective. Previous works have shown that vaccine effectiveness declines with increasing amino acid substitution distance (henceforth referred to as ‘spike sequence distance’) between circulating and vaccine variants^36,37^, with each additional 10 mutations associated with an approximately 15.4% decline in effectiveness^38^. To characterize how this distance changes over time, we analyzed over 14 million sequences collected globally between 2021 and 2025. We measured spike sequence distance between variants circulating at vaccine selection time and those observed during the following year to estimate the maximum effectiveness achievable in each period. During the first six months, 90% of observed sequences were within two mutations of the current circulating variants. By the second half of the year, this distance grew to as many as 29 mutations, which can result in a substantial drop in vaccine effectiveness and justifies annual updates. We also found that open-source fitness prediction models effectively ranked circulating variants by their future prevalence, and that a simple ranking based on current variant prevalence also performed well.

Motivated by these findings, we developed VaccineScout, a computational framework that integrates the recent genomic surveillance data with optional fitness estimates from open-source models to identify promising vaccine candidates *in silico*. VaccineScout forecasts future variant prevalence using both these quantities and recommends vaccine composition and valency (*k*), by balancing the reduction of spike sequence distances to future variants against vaccine complexity. Specifically, it identifies the point at which adding more variants yields diminishing returns. Retrospective evaluation showed that VaccineScout-recommended monovalent candidates either matched the FDA recommendations or exhibited lower spike sequence distances to future variants and higher fitness, with further gains at higher valencies. VaccineScout also approached the performance of an oracle selection with the knowledge of future variant prevalences, particularly during the initial six months after vaccine selection.

Together, these results establish VaccineScout as a systematic, *in silico* framework for guiding SARS-CoV-2 vaccine design. Specifically, VaccineScout-recommended variants can provide a more systematic and practical starting point for downstream stages of vaccine development, which include costly and time-consuming experimental characterization using neutralization and serological assays, preclinical animal studies, and human immunogenicity trials^39,40^.

## Results

### VaccineScout Overview

VaccineScout is an automated pipeline that leverages genomic surveillance data to help systematically select SARS-CoV-2 vaccine candidates and help determine their valency. It takes as input: (i) all spike sequences collected during recent genomic surveillance (default: preceding 30 days), and (ii) variant fitness estimates for each of these spike sequences from existing predictive models (optional). The framework consists of three steps: (i) forecasting future prevalence trajectories of circulating variants from their current prevalence and fitness estimates; (ii) clustering circulating variants and selecting *k* representative medoids (*k* = *1*, …, *K*, where *k* also corresponds to vaccine valency and *K* is the maximum vaccine valency the user wants to consider) that minimizes spike sequence distances between forecasted prevalence of circulating variants and their assigned medoids; and (iii) determining the optimal valency from the plot of average spike sequence distance between forecasted variants and selected vaccine candidates (Fig. 1; Methods). This average distance serves as a proxy for vaccine effectiveness against future variants, whose prevalence is unknown at the time of vaccine selection, with lower values indicating greater potential protection^36–38^. As valency (*k*) increases, the average spike sequence distance decreases, but the improvements progressively diminish, with performance eventually plateauing as additional variants provide increasingly smaller gains^41^. This curve helps determine optimal vaccine valency, beyond which the marginal reduction in spike sequence distance, and thus the marginal potential gain in protection, may no longer justify the added vaccine complexity.

**Fig. 1:**
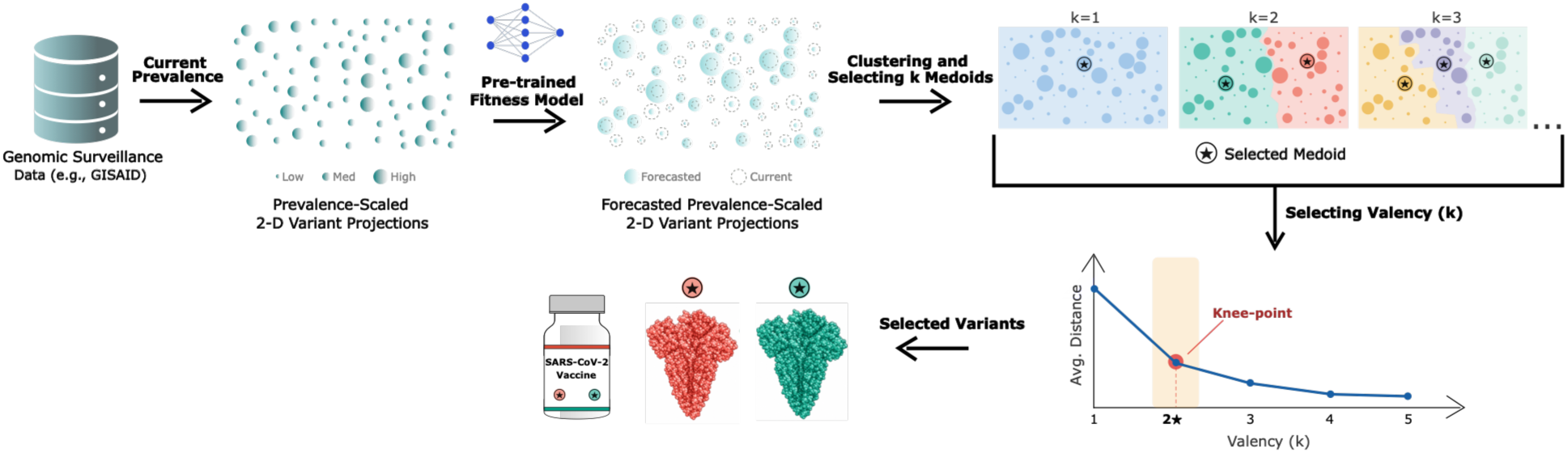
Overview of the VaccineScout pipeline. VaccineScout combines current circulating spike variant data from genomic surveillance with a pre-trained fitness predictor to recommend vaccine candidates. Circulating variants are clustered, and a representative variant (medoid) is selected for each cluster by minimizing the forecasted-prevalence-weighted spike sequence distances to all variants within the cluster. Knee-point analysis of the average spike-sequence distance (a proxy for vaccine effectiveness) versus the valency curve can be used to inform the choice of vaccine valency, which balances vaccine effectiveness with complexity. Note: the two-dimensional projections shown are for visualization purposes only and are not used by VaccineScout for clustering or vaccine candidate selection.

Variant fitness estimates from any predictive model can be incorporated into VaccineScout, including models based on protein structure, ACE2 binding affinity, DMS, antisera data, or genomic surveillance dynamics. VaccineScout also includes a tunable parameter, ***β***, that controls the time horizon for prevalence projection of current variants, with a default value of ***β*** = 60, corresponding to a forecast horizon of approximately six months (Methods). When fitness estimates are unavailable, or ***β***=0, VaccineScout assumes that future prevalence estimates remain unchanged from current levels. This allows VaccineScout to recommend vaccine candidates from genomic surveillance data alone, even when fitness estimates are unavailable.

### Currently circulating variants are responsible for a majority of infections six months in the future, and open-source fitness models are effective at forecasting dominance

We analyzed more than 14 million SARS-CoV-2 spike sequences collected globally between 2021 and 2025 to assess how long circulating variants remained prevalent. We used June 1 of each year as the cutoff date, aligning with the FDA’s annual vaccine recommendation cycle^11,42,43^, and performed the analysis for 2021–2024. For each year, we defined circulating variants using spike protein sequences collected during the 30 days preceding June 1. We then compared these sequences with those collected during two subsequent intervals: 0–6 months and 6–12 months after the cutoff date. To quantify how well circulating variants persisted into the future, we defined the ‘Normalized Tip Score’ metric. This metric calculates the fraction of sequences observed during a given future interval whose spike sequences are identical to any of the top-*k* ranked circulating variants at the cutoff date (Methods). We ranked circulating variants using three fitness prediction models: PyR_0_^32^, EVEscape^34^, and CoVFit^33^, which use different modeling approaches and data sources to estimate variant fitness. Specifically, PyR_0_ infers fitness from genomic surveillance frequency dynamics; EVEscape combines fitness estimates from a deep generative model of evolutionary sequences with structure-derived antibody accessibility and residue physicochemistry; and CoVFit fine-tunes a protein language model (ESM-2)^44^ on surveillance-derived fitness values and deep mutational scanning antibody-escape data. ESM-2’s learned representations implicitly capture structural information. We used each model’s fitness estimates to rank circulating variants and compute the Normalized Tip Score. For comparison, we also evaluated a simple circulating prevalence-based ranking and a theoretical best ranking based on each variant’s observed future prevalence. We quantified performance using the area under the curve (AUC) of Normalized Tip Score versus top-*k* ranks. Each method’s AUC was compared with the theoretical maximum AUC achieved by the perfect ranking (theoretical best ranking) of circulating variants based on observed future prevalence.

Across all years, the theoretical maximum AUC for the first six months ranged from 55–76%, indicating that variants circulating within one month of the cutoff date accounted for the majority proportion of sequences observed over the following six months (Fig. 2, Supplementary Fig. 1). All evaluated methods achieved AUCs close to the theoretical maximum (Fig. 2, Supplementary Fig. 1). CoVFit achieved the highest AUCs in most cases, with a modest advantage over PyR_0_, a larger one over EVEscape, and slightly lower performance than a naive prevalence-based ranking (mean AUC difference for PyR_0_: 0.058, EVEscape: 0.269, prevalence-based ranking: −0.019). Notably, prevalence-based ranking, which relies solely on genomic surveillance data and requires no model training, performed slightly better than more sophisticated methods, particularly during this interval. Overall, these results suggest that circulating variant prevalence and fitness estimates can both inform forecasts of future variant prevalence, providing a robust basis for selecting vaccine candidates that offer broad protection against future variants.

**Fig. 2:**
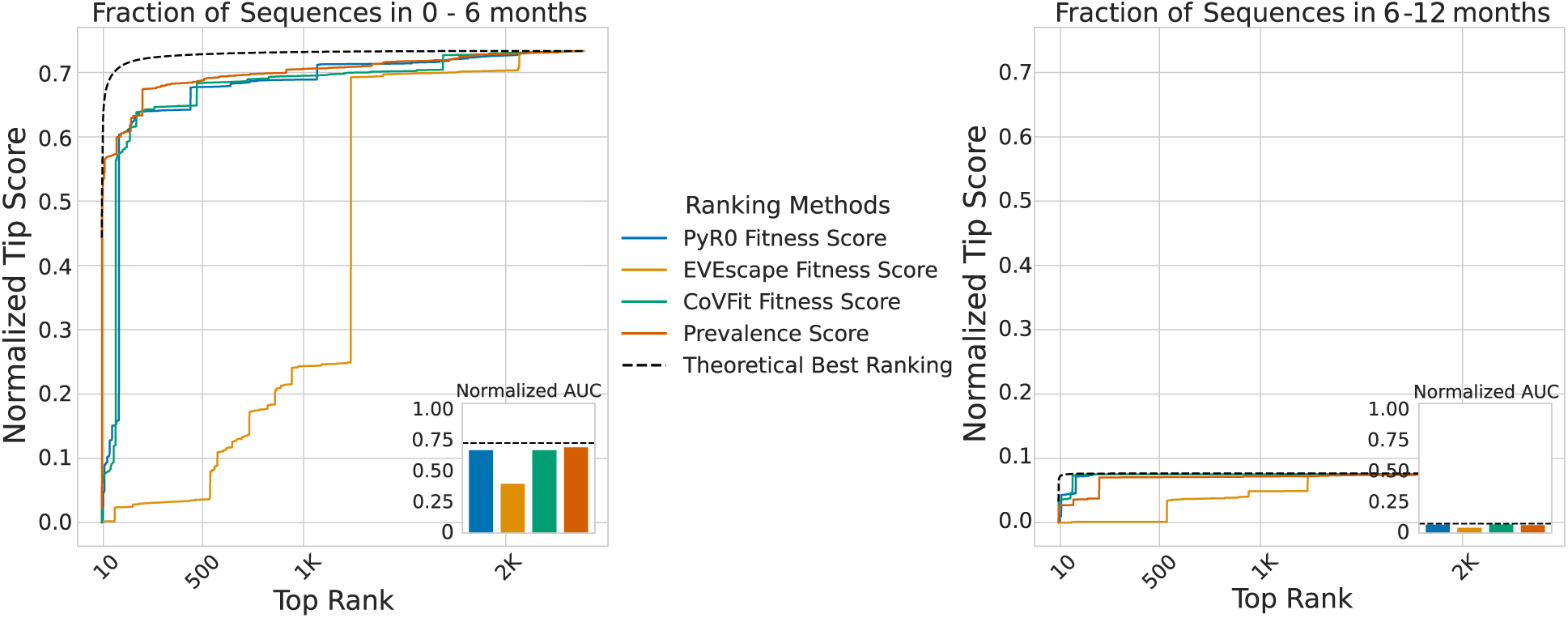
Performance comparison of fitness prediction models and prevalence-based ranking in predicting future dominance of circulating variants on the June 1, 2022 cutoff date. The line plot shows the Normalized Tip Score as a function of top-*k* ranked circulating variants predicted by PyR_0_, EVEscape, CoVFit, and Prevalence ranking models, with the Theoretical Best Ranking shown for comparison, across two future intervals: 0–6 and 6–12 months. The inset plot compares the normalized area under the curve (AUC), with a maximum value of 1, for each method with the theoretical maximum AUC for the corresponding prediction interval.

In contrast, the theoretical maximum AUC for the 6–12-month interval dropped to 5–8% across 2021–2023 and 27% in 2024. These results suggest that, beyond the initial six months, variants carrying newly acquired mutations reshape the landscape of future variants, limiting the ability of circulating variants to predict future prevalence. This motivated us to examine spike sequence divergence between circulating and future variants to assess potential vaccine effectiveness over this longer horizon.

### VaccineScout integrates variant prevalence and fitness estimates to provide a principled framework for helping determine vaccine composition

We used VaccineScout to recommend SARS-CoV-2 vaccine composition at different valencies (*k* = *1*, …, *8*) for 2021-2024, and compared these recommendations against three benchmarks: (i) FDA-recommended vaccine variants^8,11,13,14^ derived from the Mutation-Annotated Tree (MAT)^45^, (ii) the spike sequences of the clinically deployed Pfizer vaccine, and (iii) an oracle selection using the same *k*-medoids strategy as VaccineScout, but informed by the actual future prevalence of circulating variants (Methods). To evaluate potential vaccine effectiveness, we calculated the average minimum spike sequence distance between sequences collected during each future interval (0–6 and 6–12 months after vaccine selection) and the selected vaccine candidates (Fig. 3B, Supplementary Fig. 2). We also compared the fitness of selected vaccine variants using CoVFit estimates. To help determine the optimal valency, VaccineScout calculates the average minimum spike sequence distance between the forecasted prevalence of circulating variants and selected vaccine candidates at each valency. This analysis quantifies the improvement in variant coverage as valency increases (Supplementary Fig. 3).

**Fig. 3:**
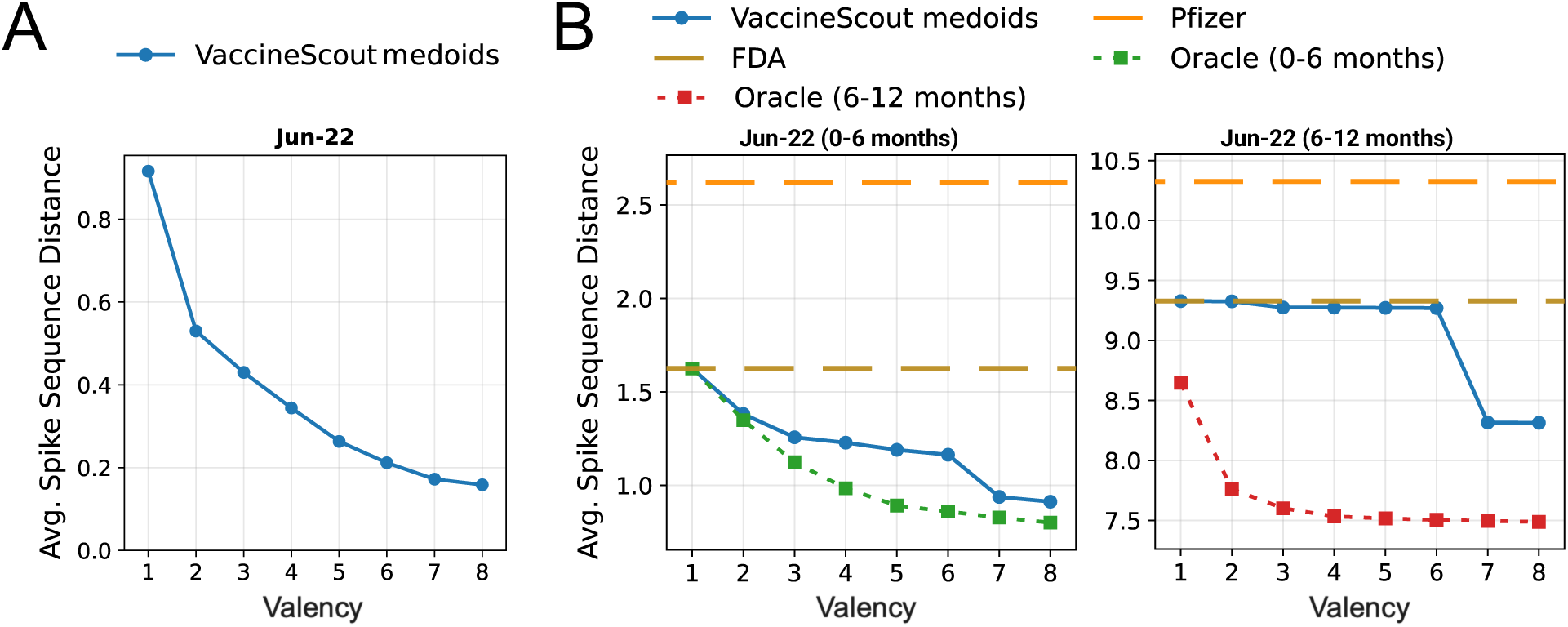
Retrospective evaluation of VaccineScout-recommended vaccine candidates across valencies. (A) Average minimum spike sequence distance of forecasted prevalence of current variants from VaccineScout-recommended candidates as a function of vaccine valency. (B) Comparison of VaccineScout-recommended, FDA-recommended, Pfizer vaccine, and oracle-selected vaccine candidates using the average minimum spike sequence distance to future variants over the 0–6 and 6–12 month intervals for the June 1, 2022 cutoff date.

From 2021 to 2024, VaccineScout’s selected monovalent variants corresponded to PANGO lineages^46^ AY.100, BA.4, EG.13, and KP.3, respectively. We compared these selections with the FDA recommendations for each year: B (the ancestral strain from Wuhan, NCBI RefSeq NC_045512.2), B + BA.4, XBB.1.5 (which has the same spike sequence as EG.13), and KP.2 (Supplementary Table 1). For the FDA’s bivalent recommendation of B+BA.4 in 2022, we considered only BA.4 in the monovalent comparison with VaccineScout, as it was more prevalent and fitter of the two variants. At the sequence level, VaccineScout’s monovalent recommendations therefore matched the FDA recommendations in 2022 and 2023, differing only in 2024. The FDA-recommended sequences were identical to the corresponding Pfizer vaccine sequences in all years except 2022 (Methods). In that year, the FDA-recommended BA.4 sequence had a higher CoVFit fitness score and a lower average minimum spike sequence distance to future variants than the Pfizer sequence (Supplementary Table 1, Supplementary Fig. 2). For 2024, VaccineScout retrospectively recommended KP.3, which differed from the FDA-recommended KP.2 by two spike mutations (T346R and Q493E). The KP.3 selection was associated with a 3.88% higher CoVFit fitness score and reduced the average minimum spike sequence distances to future variants over the first and second six-month intervals by 21.2% and 17.6%, respectively. For 2021, VaccineScout retrospectively recommended AY.100, which differed from the ancestral Wuhan sequence used in the original COVID-19 vaccine by five spike mutations (T19R, L452R, T478K, D614G, and P681R) and had a 69% higher CoVFit fitness score. Because the FDA did not recommend a circulating SARS-CoV-2 variant for vaccine strain selection in 2021, this comparison is included for completeness only. Overall, across the years evaluated, VaccineScout’s monovalent candidates either matched the corresponding FDA-recommended and Pfizer vaccine sequences or were associated with higher CoVFit fitness scores and lower average minimum spike sequence distances to future variants.

VaccineScout’s multivalent selections further reduced the average minimum spike sequence distance as valency increased, albeit with diminishing returns. In 2022, for example, the average distance during the initial six months decreased from 1.63 for the monovalent selection to 1.38 and 1.26 for the bivalent and trivalent selections, respectively, and fell by 43.8% to 0.91 for the octavalent selection. Similar trends were observed in other years (Fig. 3B, Supplementary Fig. 2A). We also evaluated VaccineScout for the more recent years 2025 and 2026, when sequencing rates had declined substantially (Supplementary Results 1). Across all years, VaccineScout’s selections remained close to the oracle selections, especially during the initial six months, with differences in average minimum spike sequence distance ranging from 0.11–0.3 across all valencies. These results indicate that fitness-based prevalence forecasting can identify vaccine variants that are close to those selected using actual future prevalence data, despite no access to future data. However, during the subsequent six-month interval (6–12 months), average minimum spike sequence distances increased substantially for both VaccineScout and oracle selections, reaching levels 10 to 20 times higher than those observed during the initial six months. This increase reflects the presence of several newly acquired mutations in future variants during this longer timeframe (Fig. 2, Supplementary Fig. 1).

As intended, the average minimum spike sequence distance between the forecasted prevalence of circulating variants and selected vaccine candidates was strongly correlated with the corresponding distance to future variants^36–38^, with the Pearson correlation coefficients ranging from 0.856 to 0.996 (Supplementary Table 2). This strong correlation supports the use of the forecast-based metric as a proxy for estimating minimum spike distance to future variants and, by extension, potential vaccine effectiveness. This metric also showed diminishing returns with increasing valency, showing a pronounced knee point at *k* = 2 across years (Fig. 3A, Supplementary Fig. 3). However, for 2021 and 2023, a trivalent vaccine was necessary to reduce the average spike distance from the forecasted prevalence of circulating variants to less than one mutation. This suggests that while a bivalent vaccine composition could offer a reasonable trade-off between improved future variant coverage and vaccine design complexity and costs, a more systematic analysis under an explicit design cost model from vaccine manufacturers could help identify the optimal balance.

### Future dominant SARS-CoV-2 variants are substantially mutated from circulating variants beyond six months, but most mutations are nearly neutral, limiting the ability to predict future mutations

Average minimum spike sequence distances to future variants were substantially higher during the 6–12-month interval than during the initial six months, even for the oracle-selected candidates, suggesting a potential rapid decline in vaccine effectiveness over this period^38,47,48^ (Supplementary Fig. 2B). To investigate this trend, we examined the duration of dominance of different SARS-CoV-2 PANGO lineages, defined by their spike sequences, among sequences collected between May 2021 and May 2025. The duration of prevalence varied considerably, ranging from approximately eleven months for KP.3 to just three months for B.1.1.7, BA.1, BA.2.12.1, and HV.1 before these lineages were displaced by others (Fig. 4A). A closer examination of two representative selection years, 2021 and 2024, illustrates how lineage turnover can shape the potential effectiveness of vaccine candidates over time.

**Fig. 4:**
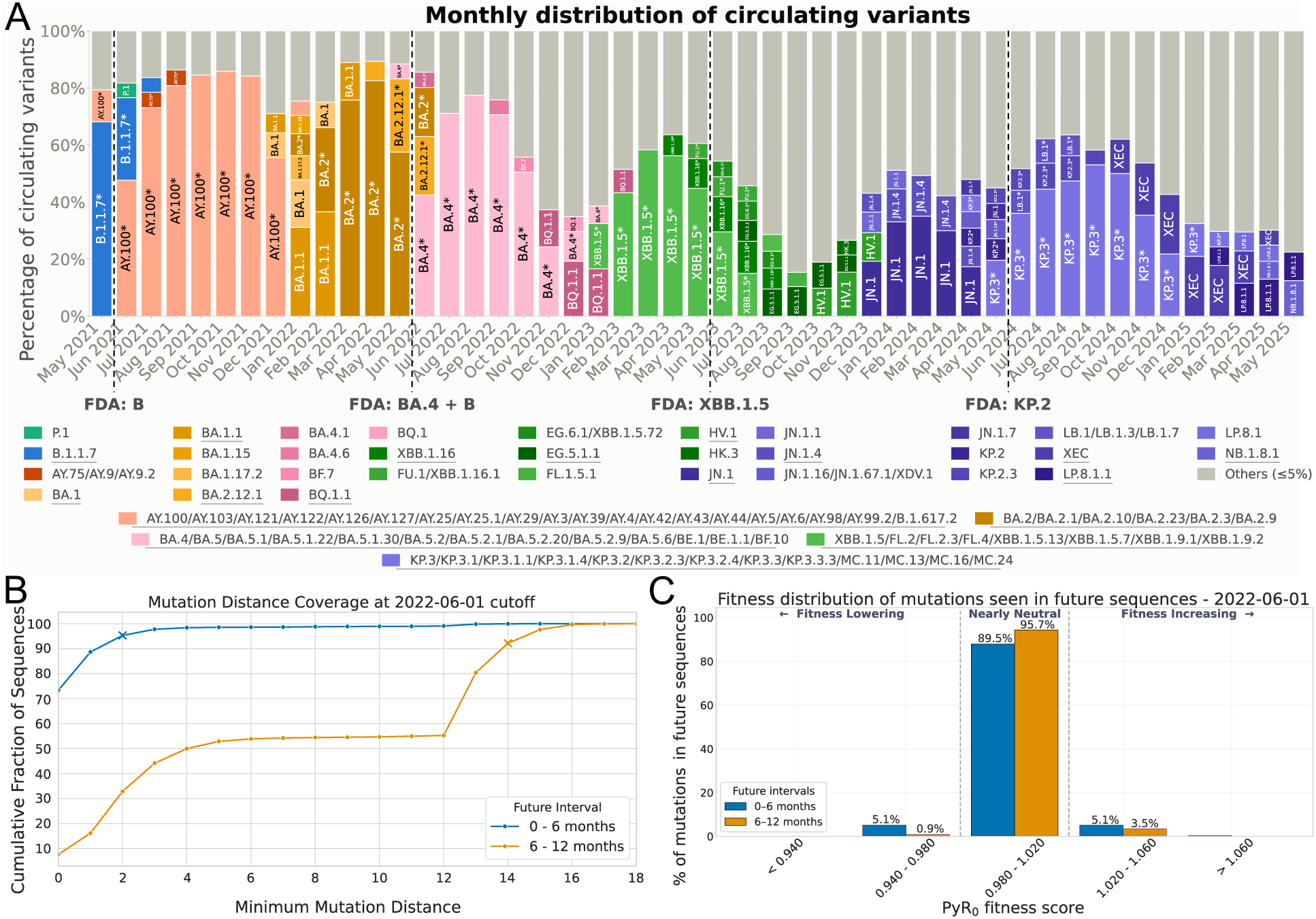
Monthly distribution of circulating variants from May 2021 to May 2025, and distribution of acquired mutations in future variants across the 0-6- and 6-12-month intervals following the June 1, 2022 cutoff date. (A) Circulating variants emerging and becoming dominant over time, with variants exceeding 5% prevalence highlighted and those exceeding 10% underlined in the legend. Variants with identical spike sequence are clubbed together. The dotted lines mark the months when the past FDA recommendations were issued, with the corresponding variants labelled. (B) Distribution of minimum spike sequence distance of future variants relative to all circulating variants, with ‘x’ denoting the points on the two curves reaching 90% of cumulative sequences. (C) Distribution of PyR_0_-estimated relative fitness scores for newly acquired mutations in future variants.

At the 2021 selection time, B.1.1.7 (Alpha) and AY.100 (Delta) were the two major lineages, with AY.100 remaining dominant for approximately seven months before being completely displaced by Omicron (BA.1 and BA.1.1) in January 2022 (Fig. 4A). VaccineScout’s monovalent recommendation of AY.100 therefore corresponded to the dominant circulating lineage through the end of 2021, indicating that the recommended vaccine provided a close match to circulating variants during this period. However, this similarity would have been eroded with the emergence of Omicron, whose BA.1 and BA.1.1 spike sequences differed from AY.100 by 28 and 29 mutations, respectively. In 2024, five significant lineages were co-circulating during vaccine selection time: KP.3, KP.2, KP.2.3, JN.1, and JN.1.16. The FDA initially recommended JN.1 before revising its recommendation to KP.2 a week later. In contrast, VaccineScout’s monovalent recommendation, KP.3, remained at a significant proportion through February 2025, considerably longer than the FDA-recommended KP.2 (Fig. 4A). Notably, at the time of selection, KP.3 differed from the co-circulating lineages by only 2–3 spike mutations. The subsequent XEC, LP.8.1, and LP.8.1.1 lineages that displaced KP.3 differed from it by 2–6 mutations, whereas the corresponding distances from the FDA-selected KP.2 were 4-6 mutations. To quantify this genetic divergence, we calculated minimum spike sequence distance between each future variant and any circulating variant at vaccine selection time across the 0–6 and 6–12-month intervals (Fig. 4B). During the initial six months, 90% of future sequences accumulated two or fewer mutations relative to circulating variants. During the subsequent six months, however, the number of mutations increased substantially, reaching 14 to 29 in 2021–2023 and six in 2024, which could be an artifact of the smaller number of sequences collected in that year (Supplementary Fig. 4). This substantial mutation accumulation in future variants explains the elevated spike sequence distances observed for both VaccineScout- and oracle-selected vaccine candidates during the 6–12-month interval, highlighting an inherent limitation of selecting vaccine candidates solely from currently circulating variants when viral evolution can substantially alter the spike sequence landscape over time.

Spike sequence distances beyond six months might potentially be further lowered if mutations that will arise in future variants could be predicted in advance and incorporated into vaccine candidates. To assess the feasibility of this approach, we examined the fitness effects of mutations observed in future sequences. Using PyR_0_, we classified each acquired mutation as fitness-lowering (<0.98), nearly neutral (0.98-1.02), or fitness-increasing (>=1.02) across the 0–6- and 6–12-month intervals. In both intervals, the majority of acquired mutations were nearly neutral, accounting for 82.1–92.8% of mutations during 0-6 months and 55.8%–95.7% during 6-12 months. Fitness-increasing mutations were relatively less frequent, although their proportion increased from 3.5% to 43.8% during the one year after cutoff across 2021-2023. The largest increase was observed in 2021 coinciding with the Omicron emergence (Fig. 4C, Supplementary Fig. 5). The fitness distribution of mutation events inferred from the phylogenetic tree as future variants emerged indicates that most acquired mutations are likely ‘passengers’ that arise alongside a smaller number of fitness-increasing ‘driver’ mutations that promote the spread of the resulting variants through the population^49,50^ (Supplementary Fig. 6). These findings are consistent with many prior works^51,52^, including Ohta’s ‘nearly neutral theory of molecular evolution’, which posits that most mutations reach fixation by chance through genetic drift acting on neutral or nearly neutral mutations, whereas only a small fraction confer substantial selective advantages^53^. Because the space of nearly neutral mutations is vast, and their fixation is largely stochastic, predicting which ones will occur and become prevalent in future variants therefore remains inherently challenging. These findings are consistent with the limited success of previous efforts in predicting mutations: although such methods can forecast the near-term spread of mutations already observed in circulating variants^54^, predicting the new mutations acquired by future variants remains an unsolved problem^54,55^. Consequently, we posit that vaccine design strategies that rely on predicting future mutations may offer limited advantage over selection directly from circulating variants.

## Discussion

Vaccination remains an essential strategy for reducing SARS-CoV-2 infections and disease burden amid ongoing viral evolution. However, rapid and inexpensive *in silico* screening methods to identify strong vaccine candidates for prioritization in costly, time-consuming downstream experimental validation remain lacking. VaccineScout fills this gap by formalizing vaccine design as an optimization problem, specifically, as one that minimizes the average minimum spike sequence distance between selected variants and the prevalence of circulating variants in the future. Despite its simplicity, VaccineScout recommendations closely matched the final FDA recommendations that were informed by extensive experimental data, and even outperformed them on certain metrics. These findings highlight VaccineScout’s potential as an efficient and principled data-driven framework for early vaccine candidate screening and prioritization for downstream experimental validation.

Importantly, VaccineScout relies on genomic surveillance data and open-source variant fitness models, avoiding dependence on proprietary serological data currently used in vaccine design and enabling rapid adaptation to changing epidemiological trends. Although trained fitness models provide additional information about variant properties, including information about protein structure, DMS, and antisera measurements, we showed that VaccineScout also performed well when using genomic surveillance data alone. This flexibility further supports its utility during the early stages of an outbreak or pandemic, when such models and data may not yet be available. This makes VaccineScout complementary to the approach of Nasir et al.^17^, which uses serum neutralization data to predict immune escape and antigenic relationships among emerging variants to guide vaccine variant selection. VaccineScout could be further extended by incorporating immune escape predictions from their model, enabling vaccine candidates to be selected with the aim of improving protection against future variants.

Besides the computational framework, this work provides a comprehensive analysis of global SARS-CoV-2 genomic surveillance data, yielding valuable insights for vaccine updates. Our findings provide quantitative support for the need for annual vaccine updates, as the average spike sequence distance between selected vaccine candidates and circulating variants increases substantially between 6 and 12 months of selection, potentially reducing vaccine effectiveness. At the same time, our results demonstrate that selecting vaccine candidates from currently circulating variants remains an effective strategy for providing population-level protection against ongoing viral evolution. We further found that newly acquired mutations that eventually become fixated in the population are difficult to predict, due to the inherently stochastic nature of viral evolution. Hence, vaccine design strategies aimed at improving vaccine effectiveness by predicting future mutations may offer limited benefits.

In terms of limitations, VaccineScout currently considers only amino acid substitutions in the SARS-CoV-2 spike protein when minimizing distances among circulating variants. Incorporating insertions and deletions (indels) could improve candidate selection by capturing additional biologically relevant variation. However, accounting for indels complicates distance computation because their variable lengths make it challenging to determine appropriate weights relative to individual substitutions^56,57^. Nonetheless, our existing substitution-based approach remains well justified because substitutions greatly outnumber indels across SARS-CoV-2 lineages and are strongly correlated with them^58–60^. One possible extension would be to compute distances in the embedding space of spike sequences generated by a protein language model, such as ESM-2^44^. This approach would preserve VaccineScout’s distance-based formulation while implicitly capturing indels and other forms of sequence variations^61–63^. Secondly, as VaccineScout selects vaccine candidates from variants observed through genomic surveillance, its performance depends on genomic surveillance coverage and on how early emerging variants can be detected. Retrospective analysis for 2025 and 2026 nevertheless showed that VaccineScout-recommended candidates achieved lower spike sequence distance to future variants than FDA recommendations, demonstrating its potential even during limited genomic surveillance (Supplementary Results 1). If clinical genomic surveillance further declines, one possible strategy to improve VaccineScout’s performance would be through incorporating wastewater surveillance data, which can detect emerging and novel variants weeks before they are detected through clinical genomic surveillance^60,64^.

Overall, VaccineScout provides a systematic framework for prioritizing candidate vaccine variants by projecting future prevalence from recent genomic surveillance data and fitness estimates. By accounting for projected evolutionary trajectories rather than solely on currently circulating variants, VaccineScout’s recommended candidates had greater spike sequence similarity to future variants than previous FDA recommendations across the years, potentially improving vaccine effectiveness while enabling more timely adaptation to changing viral dynamics^36–38^. More broadly, this work provides a systematic data-driven approach to incorporating evolving genomic surveillance data into vaccine candidate selection, with potential applicability to other rapidly evolving pathogens with sufficient ongoing genomic surveillance data.

## Methods

### SARS-CoV-2 spike and vaccine sequence data

#### Sequence data

We extracted SARS-CoV-2 genome sequences from mutation-annotated trees (MATs) maintained using UShER^45,65^. These MATs provide a comprehensive phylogenetic representation of all available SARS-CoV-2 sequences from GenBank^66^, COG-UK^67^, and the China National Center for Bioinformatics^68^, as well as restricted-access sequences from GISAID^69^, and include PANGO lineage annotations^46^ and inferred mutations for each node. We used the MAT dated September 1, 2025, for most analyses. For VaccineScout recommendations for years 2025 and 2026 and their comparisons with the corresponding FDA recommendations, we instead used the MAT dated September 1, 2026. From these MATs, we also extracted the spike sequences of over 14 million SARS-CoV-2 sequences collected between 2021 and 2025.

#### Sequence dating

We used Chronumental^70^ to estimate the dates of all nodes in both MATs. For each year from 2021 to 2024, we defined circulating sequences as those collected during the 30 days preceding the June 1 cutoff date. To obtain circulating variants, we removed duplicate sequences and excluded variants represented by fewer than five sequences to mitigate the potential effects of date mislabeling in public databases. We applied the same deduplication procedure to sequences collected during the first six months after each cutoff and the subsequent 6–12-month interval, using the resulting unique variants to obtain future variant prevalence in each period.

#### FDA-recommended and Pfizer vaccine composition data

FDA-recommended vaccine variants for each year^8,11,13,14^ were extracted from the MAT by identifying the annotated node corresponding to each variant and reconstructing its nucleotide sequence from the mutations along the path from the root to that node. We then translated these mutations into amino acid substitutions and retained only those within the spike protein region to obtain the corresponding spike sequences. Pfizer vaccine sequences for Tozinameran (2021–2022), Famtozinameran and Tozinameran (2022–2023), Raxtozinameran (2023–2024), and Cemivameran (2024–2025) were downloaded from https://drugs.ncats.io. To ensure a consistent comparison between these Pfizer spike sequences, VaccineScout candidates, and MAT-extracted FDA-recommended variants, we made the following changes to the Pfizer spike sequences: (i) removed any insertions or deletions, as the MAT represents only substitutions; (ii) masked sites 986 and 987, which contain prefusion-stabilized SARS-CoV-2 spike mutations^71^; and (iii) masked sites 24 and 211, which are subject to branch-specific masking for variants under consideration in the MAT, and site 142, which is masked across the entire MAT. After these changes, all Pfizer spike sequences matched their corresponding MAT-derived FDA variant exactly, except Famtozinameran, which retained a single mutation at site 408 (Supplementary Fig. 2).

### Evaluation Metrics and Comparison Methods

We used the following metrics to evaluate the performance of fitness model- and prevalence-based circulating variant rankings and to compare VaccineScout-recommended vaccine candidates with FDA recommendations based on their spike sequence similarity to future variants.

- **Normalized Tip Score** - Fraction of future sequences collected during a six-month interval (0-6 months and 6-12 months) after the June 1 cutoff, whose spike sequences are identical to any of the top-*k* ranked circulating variants collected during the 30 days preceding the cutoff. Circulating variants were ranked using either fitness estimate models from PyR_0_, EVEscape, and CoVFit, or a circulating prevalence-based ranking model.
- **Area under the curve (AUC) of Normalized Tip Score versus top-*k* rank** - Normalized Tip Score is computed for each *k* rank from 1 to the total number of circulating variants, and the resulting values are used to calculate the area under the curve (AUC) using the trapezoidal rule. The AUC is normalized by the total number of circulating variants to account for differences in the number of circulating variants across different years, yielding a value between 0 and 1.
- **Prevalence-based ranking** - Circulating variants collected during the 30 days preceding the June 1 cutoff date were ranked according to the frequency of sequences observed for each variant, with variants having higher observed prevalence ranked higher.
- **Theoretical best ranking** - Circulating variants are ranked based on the frequency of their corresponding spike sequences observed during the corresponding six-month future interval (0-6 months and 6-12 months).
- **Average minimum spike sequence distance from future sequences** - Spike sequence distance (Dist) is defined as the number of amino acid sites that differ between two spike sequences. For a set of *m* future sequences {f_i_}and *n* selected vaccine sequences {s_j_}, the average minimum spike sequence distance is defined as:

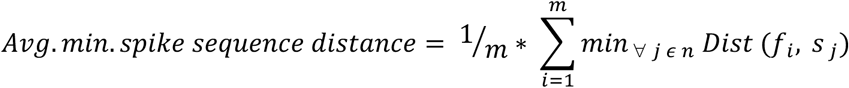
- **Average minimum spike sequence distance from forecasted prevalence of circulating variants** - This metric is used to inform the choice of vaccine valency by first forecasting the prevalence of circulating variants using their circulating prevalence and fitness estimates, and then using this forecasted prevalence to weigh the average minimum spike sequence distance of circulating variants to vaccine candidates. This distance shows a strong correlation with the average minimum spike sequence distance to future variants (Supplementary Table 2). For a set of *m* circulating variants {c_i_}, with corresponding prevalence values {p_i_}and fitness estimates {r_i_}, and *n* selected vaccine sequences {s_j_}, the forecasted prevalence is first calculated as:

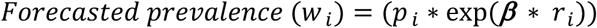

where *β* is a tunable parameter controlling the time horizon of prevalence projection. The default value of ***β*** = 60 corresponds to a forecast horizon of approximately six months (Supplementary Fig. 7). Increasing the value of *β* extends the fitness-based prevalence forecast further into the future by assigning greater weight to variants with higher fitness. The formulation is robust to the choice of *β*, with comparable performance observed across a range of values in different years and for different medoid selections. The average weighted spike sequence distance is then defined as:

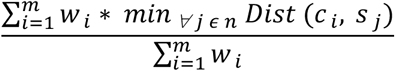

### VaccineScout variant selection

VaccineScout formulates vaccine composition selection as a weighted *k*-medoids optimization problem^72^ over circulating SARS-CoV-2 spike sequences. Given *m* circulating variants, each with observed prevalence and fitness estimate, VaccineScout selects *n* representative vaccine variants (medoids) that minimize the spike sequence distance from the forecasted prevalence of circulating variants, as described above. In the current implementation, prevalence forecasting uses CoVFit fitness estimates, which incorporate surveillance-derived fitness and deep mutational scanning data on antibody escape through a protein language model that captures structural information. Thus, although spike sequence distance assigns equal weight to each amino acid difference, its contribution is weighted by the forecasted prevalence of each variant, which is informed by these fitness estimates and their underlying data sources. Consequently, the contribution of the same sequence distance to vaccine candidate selection varies depending on the fitness effects of the mutations in the corresponding variant. Each circulating variant is assigned to its closest selected medoid, with variants having higher projected prevalence contributing more strongly to medoid selection. As additional variants are included in the vaccine composition, this spike sequence distance from forecasted prevalence of variants decreases, providing a metric for estimating sequence coverage of future variants (Supplementary Fig. 3).

However, the magnitude of this distance reduction diminishes with increasing valency. We therefore select the vaccine valency at the knee point of the distance versus valency curve, where additional vaccine variants provide diminishing returns.

### Oracle variant selection

Oracle vaccine variant selection provides a theoretical upper bound on vaccine selection performance by using the observed prevalence of variants during each future six-month interval (0–6 months and 6–12 months) following the June 1 cutoff date. For each interval, the oracle selects *n* vaccine candidates from among the variants circulating at the cutoff date. Unlike VaccineScout, which forecasts the prevalence of circulating variants in the future using fitness estimates, the oracle uses the actual observed future prevalence, which is not available at the cutoff date, thus representing optimal vaccine composition.

### Analyzing mutations with PyR0

PyR_0_ amino acid scores were obtained from the ‘mutations.tsv’ file in the January 4, 2024 model output available in the Broad Institute *pyro-cov* GitHub repository: https://github.com/broadinstitute/pyro-cov. The original file contained fitness scores for 3,792 amino acid mutations. To expand the set of ranked mutations, we additionally inferred scores for reversal and chain mutations that were absent from the original output. For reversal mutations, if the score for S:A10G was available but that for S:G10A was not, the latter was assigned a score of 1/PyR_0_(S:A10G). For chain mutations, if scores for S:A10G and S:G10N were available but S:A10N was not, the latter was assigned a score of PyR_0_(S:A10G) * PyR_0_(S:G10N).

For ranking circulating variants, the PyR_0_ score of each variant was calculated as the product of the ‘R/R_A’ scores of its constituent amino acid mutations present in the expanded mutation set described above (Supplementary Fig. 1). For any spike sequence mutation not present in the expanded mutation set, we assigned it a fitness score of 1.0.

The fitness distribution of mutations acquired by future sequences in the 0–6- and 6–12-month intervals was determined by accumulating mutations along the path from the internal node dated at or immediately before the cutoff date to each sequence observed within the corresponding interval and plotting their PyR_0_ fitness scores as a histogram. Mutations with scores <0.98 were classified as fitness-lowering, those with scores from 0.98 to 1.02 as nearly neutral, and those with scores ≥1.02 as fitness-increasing (Supplementary Fig. 5). Separately, the fitness distribution of mutation events observed in the phylogenetic tree as future variants emerged in each interval was computed by tracing the path from an internal node dated at or immediately before the cutoff date to each sequence (leaf node) observed within the corresponding interval. Each path was traversed only once, while accounting for all combinations of mutations observed at the leaf nodes along that path (Supplementary Fig. 6).

### Scoring variants with EVEscape

For ranking circulating variants using EVEscape, we selected a reference variant for calculating EVEscape scores (https://evescape.org/data) based on the FDA-recommended vaccine variant from the preceding year: Wuhan for 2021 and 2022, BA.4 for 2023, and XBB for 2024. For each year, the EVEscape score of a circulating variant was calculated as the sum of the ‘EVEscape’ scores of its constituent amino acid mutations relative to the corresponding reference variant (Supplementary Fig. 1).

### Scoring variants with CoVFit

For ranking circulating variants using CoVFit, we downloaded the CoVFit-CLI from https://zenodo.org/records/14438178. Fitness scores were obtained by providing a FASTA file containing the spike sequences of the circulating variants as input to CoVFit-CLI using the following command: “./covfit_cli -i {fasta_path} -o {covfit_outdir} -g -b 16 -f 3”. We used the ‘fitness_mean’ values reported in the resulting output file as the CoVFit fitness scores for ranking circulating variants.

## Data Availability

The VaccineScout source code used in this study is freely available under the MIT license at https://github.com/TurakhiaLab/VaccineScout. All analyses in this study were performed using publicly available SARS-CoV-2 sequences obtained from a mutation-annotated tree and other publicly available data, with their respective sources described in the Methods section. All manuscript figures representing different analyses are provided as Jupyter notebooks, while VaccineScout itself is a standalone Python command-line tool that users can run on their own data.

## Code and data availability

The VaccineScout source code used in this study is freely available under the MIT license at https://github.com/TurakhiaLab/VaccineScout. All analyses in this study were performed using publicly available SARS-CoV-2 sequences obtained from a mutation-annotated tree and other publicly available data, with their respective sources described in the ‘Methods’ section.

All manuscript figures representing different analyses are provided as Jupyter notebooks, while VaccineScout itself is a standalone Python command-line tool that users can run on their own data. VaccineScout requires the UShER mutation-annotated tree (MAT) for genomic surveillance data, the Wuhan reference sequence, and the CoVFit CLI for variant fitness estimation, with optional metadata containing SARS-CoV-2 sequence collection dates. For each valency *k* (*k* = 1 to 8 by default), VaccineScout generates a CSV containing the selected vaccine candidates, their CoVFit fitness estimates, PANGO lineages^46^, and the average minimum spike-sequence distance between the forecasted prevalence of circulating variants and the selected candidates, along with a FASTA file containing the spike sequence of the selected vaccine candidates.

## Acknowledgements

We gratefully acknowledge all data contributors—namely, the authors and their originating laboratories responsible for specimen collection, and the submitting laboratories for generating and sharing sequence data and metadata via GISAID, GenBank, COG-UK, and the China National Center for Bioinformation— on which this research is based. This work was supported by funding from the U.S. Centers for Disease Control and Prevention through the Office of Advanced Molecular Detection (CDC contracts #75D30123C17463 and #75D30124C20302), the Amazon Research Award (Fall 2022 CFP), and the National Artificial Intelligence Research Resource, NAIRR240477.

## Supplementary Material

### Supplementary Results 1: Evaluating VaccineScout recommendations for 2025 and 2026

We compared VaccineScout candidates against the FDA recommendations for 2025 and 2026 using a September 1, 2026 MAT and the same CoVFit model used in the earlier analyses, dated October 7, 2024. Evaluation was challenging because sequencing rates have declined substantially in recent years. In the 30 days before the June 1 cutoff, we identified 15,322 sequences representing 2,632 variants in 2025 and 2,667 sequences representing 685 variants in 2026. After filtering out variants represented by fewer than five sequences, only 272 and 76 variants remained, respectively.

The FDA recommended variants LP.8.1 for 2025 and XFG for 2026, while VaccineScout selected monovalent candidates, MC.33.1 and RC.1.1, respectively. For 2025, the average minimum spike sequence distances to future variants were 9.65 for LP.8.1 and 8.47 for MC.33.1 during the first six months after selection, and 15.76 and 14.07, respectively, during the subsequent six months. VaccineScout’s trivalent candidates reduced these distances to 1.46 and 7.64, respectively, while its octavalent candidates further reduced them to 1.16 and 7.20. For 2026, the distances to variants observed during the three months after selection were 14.83 for XFG and 13.36 for RC.1.1, decreasing to 6.96 with VaccineScout’s octavalent selection.

LP.8.1 and MC.33.1 differed by three spike mutations (L186F, S190R, and R1086K), with CoVFit scores of 0.8554 and 0.8549, respectively. XFG and RC.1.1 differed by 12 mutations (P31S, F59S, R182K, G184S, S190R, T346R, A435S, R444K, R445P, K478T, D487N, and I572T), with scores of 0.8523 and 0.8458, respectively. VaccineScout’s monovalent candidates had slightly lower CoVFit scores in both years, potentially reflecting the use of an older model. Nevertheless, this model performed well in forecasting future variant prevalence, enabling VaccineScout to select candidates with lower distances to future variants than the corresponding FDA recommendations for both years, even with limited genomic surveillance data.

### Supplementary Figures

**Supplementary Fig. 1:**
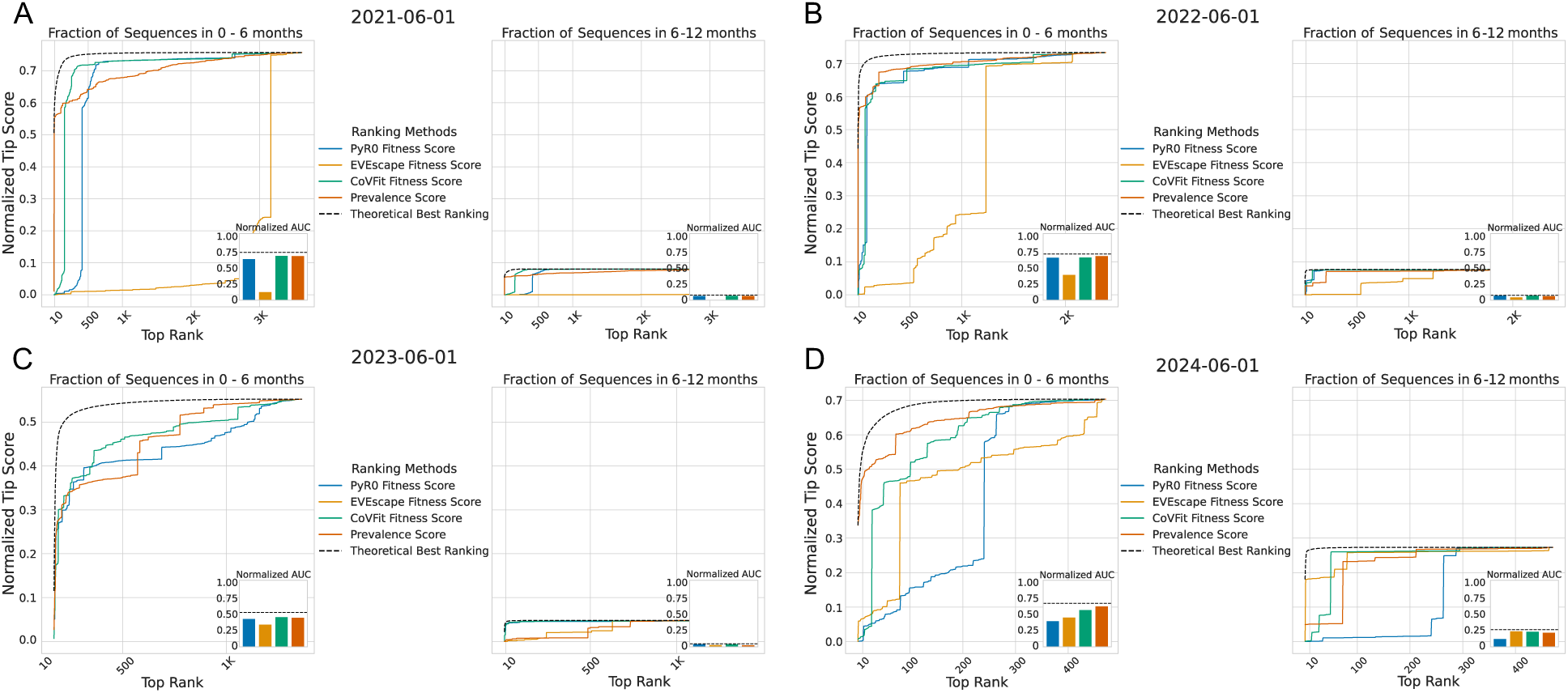
Performance comparison of fitness prediction models and prevalence-based ranking in predicting future dominance of circulating variants at the June 1 cutoff date for (A) 2021, (B) 2022, (C) 2023, and (D) 2024. The line plot shows the Normalized Tip Score as a function of top-k ranked circulating variants predicted by PyR_0_, EVEscape, CoVFit, and Prevalence ranking models, with the Theoretical Best Ranking shown for comparison, across two future intervals: 0–6 and 6–12 months. The inset plot compares the normalized area under the curve (AUC), with a maximum value of 1, for each method with the theoretical maximum AUC for the corresponding prediction interval.

**Supplementary Fig. 2:**
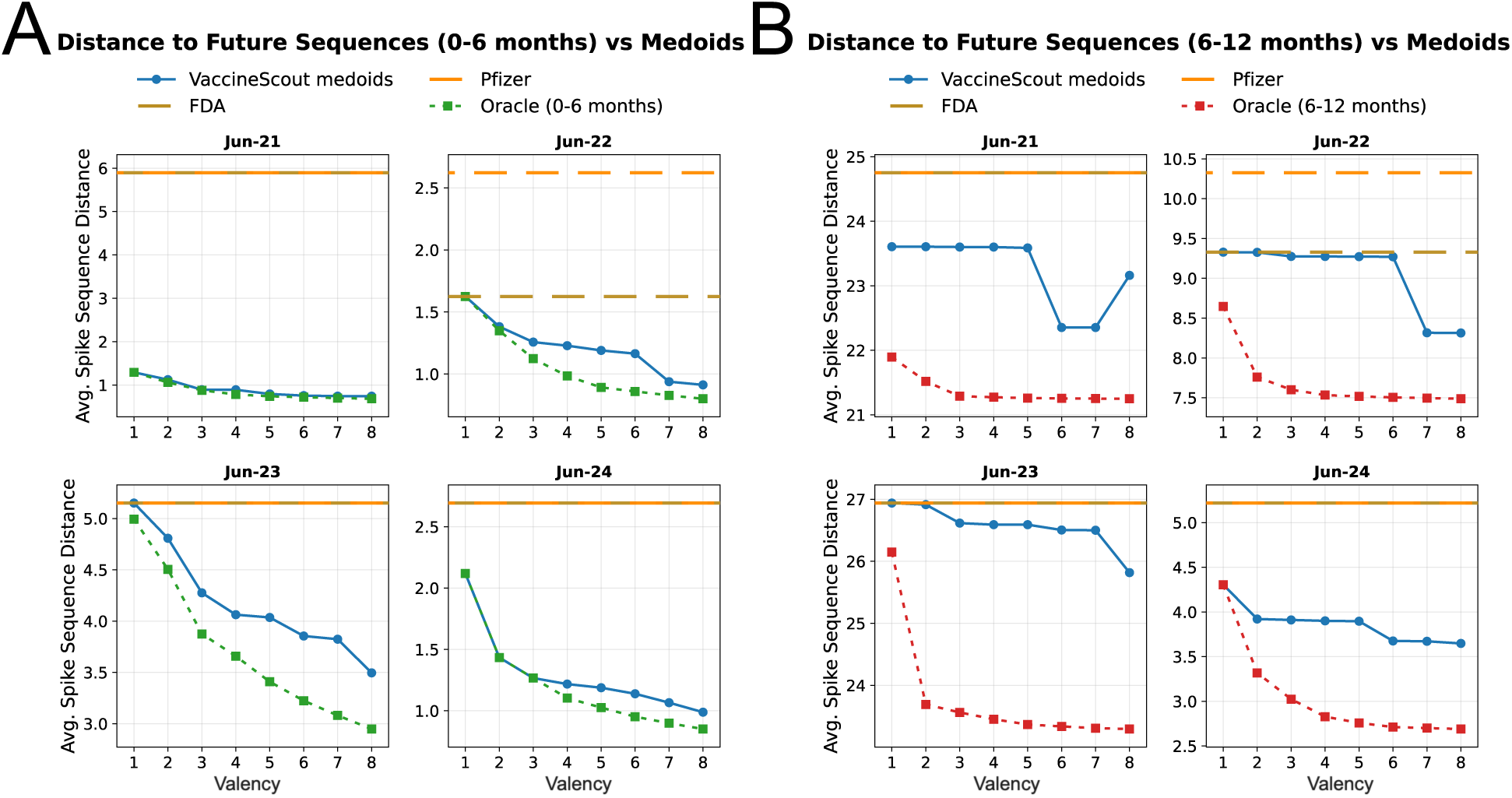
Comparison of VaccineScout-selected vaccine candidates across valencies against FDA-recommended (MAT-derived), Pfizer vaccine, and oracle-selected vaccine candidates using the prevalence-weighted average minimum spike sequence distance to future variants over the (A) 0–6 month and (B) 6–12 month intervals for the June 1 cutoff date across years 2021-2024.

**Supplementary Fig. 3:**
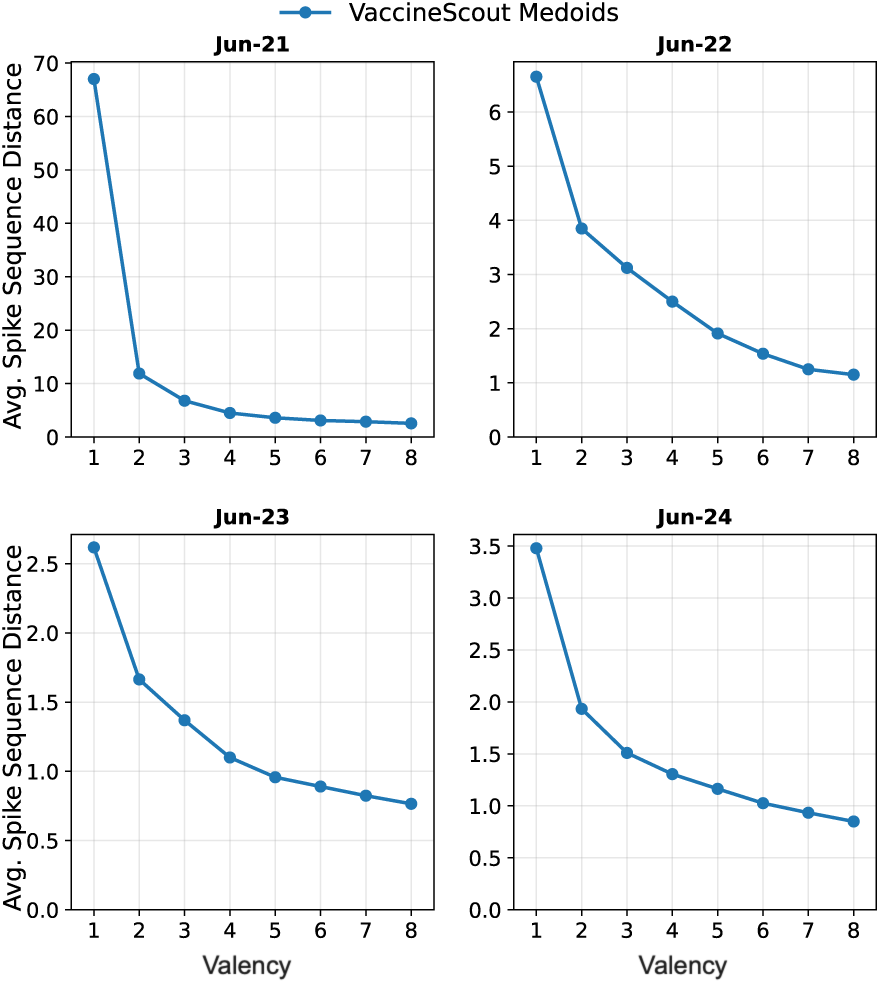
Average spike sequence distance between the forecasted prevalence of circulating variants and VaccineScout-selected candidates as a function of vaccine valency at the June 1 cutoff date across 2021-2024.

**Supplementary Fig. 4:**
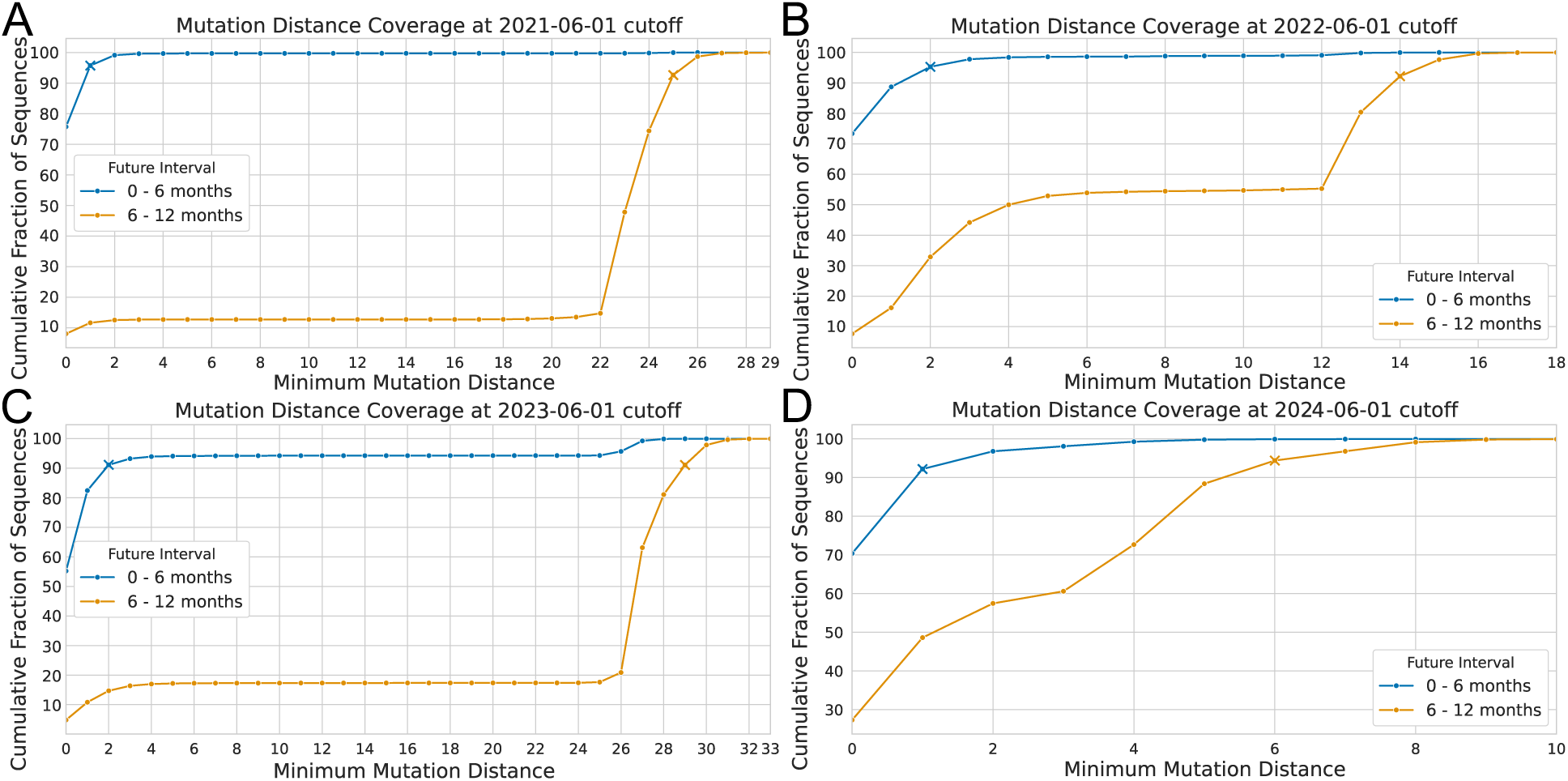
Number of acquired mutations by future variants across the 0-6- and 6-12-month intervals following the June 1 cutoff date for (A) 2021, (B) 2022, (C) 2023, and (D) 2024. ‘x’ denotes the points on the two curves reaching 90% of cumulative sequences.

**Supplementary Fig. 5:**
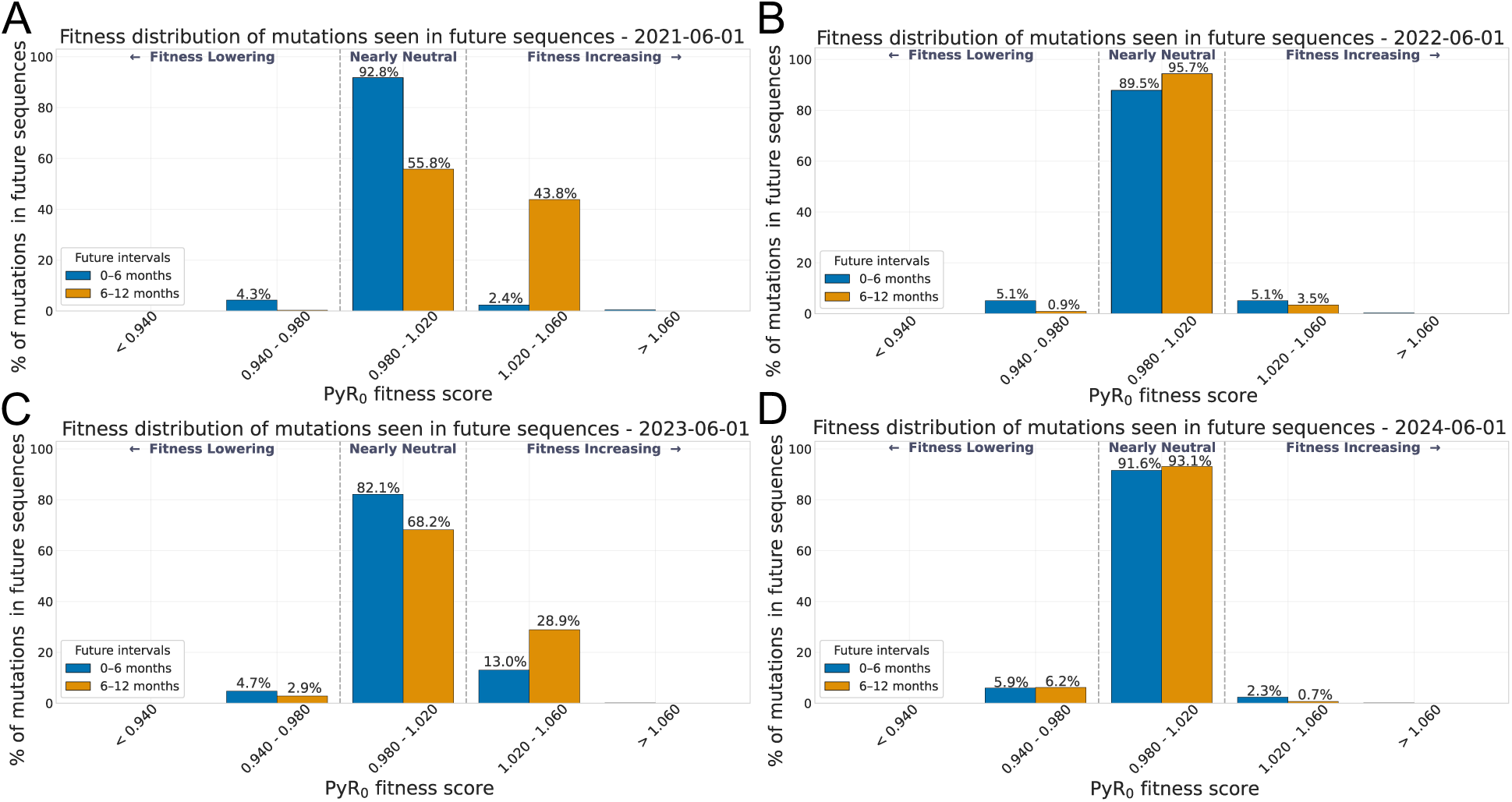
Fitness distribution of mutations acquired by future variants, determined using PyR_0_ across the 0-6- and 6-12-month intervals following the June 1 cutoff date for (A) 2021, (B) 2022, (C) 2023, and (D) 2024.

**Supplementary Fig. 6:**
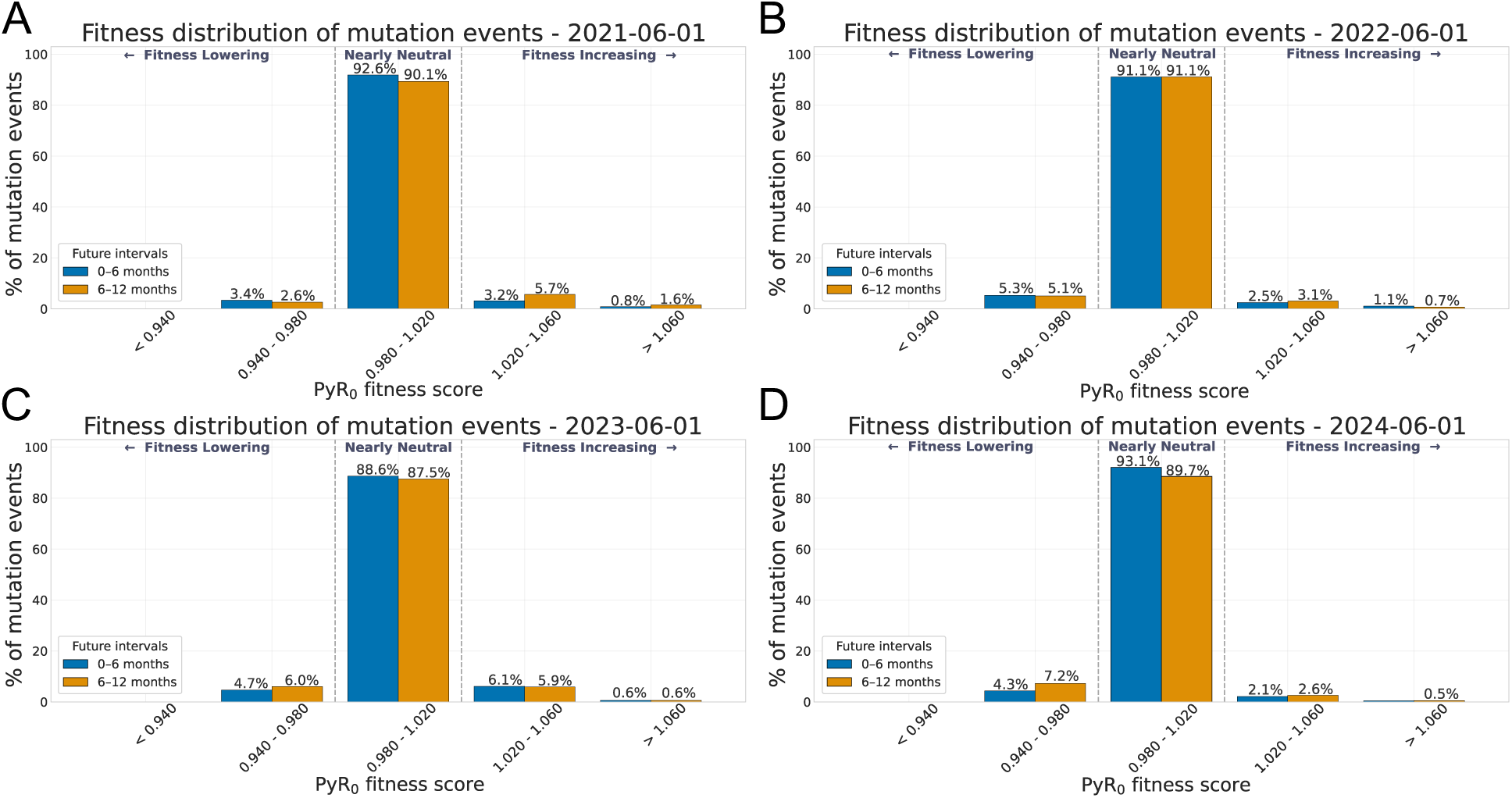
Fitness distribution of mutation events observed in the phylogenetic tree with emergence of future variants, determined using PyR_0_ across the 0-6- and 6-12-month intervals following the June 1 cutoff date for (A) 2021, (B) 2022, (C) 2023, and (D) 2024.

**Supplementary Fig. 7:**
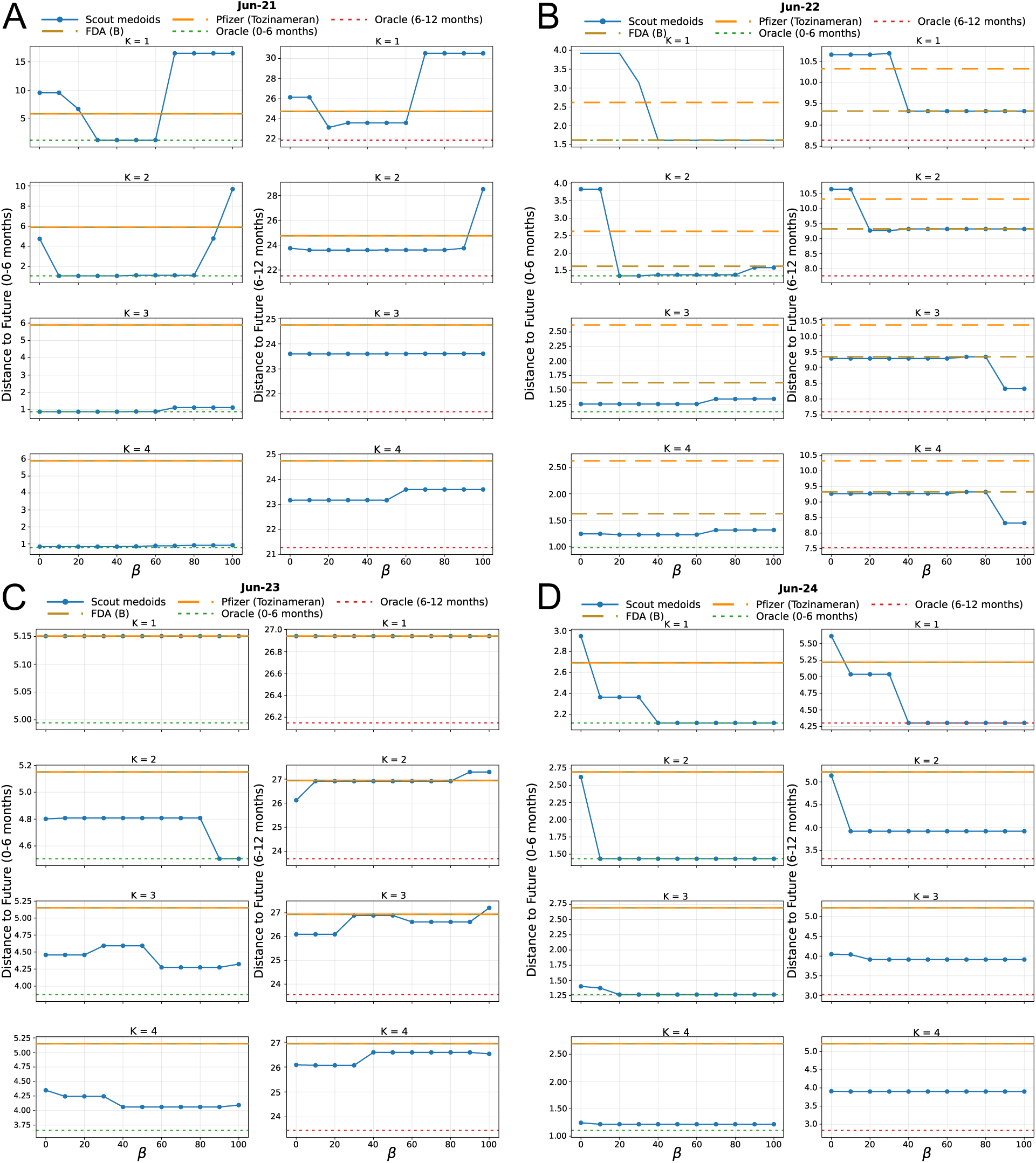
Comparison of VaccineScout’s selected variants using different values of *β* in prevalence-weighted average minimum spike sequence distance to future variants over the 0–6-month and 6– 12-month intervals following the June 1 cutoff date for (A) 2021, (B) 2022, (C) 2023, and (D) 2024.

